# Post-Acute Cardiovascular Outcomes of COVID-19 in Children and Adolescents: An EHR Cohort Study from the RECOVER Project

**DOI:** 10.1101/2024.05.14.24307380

**Authors:** Bingyu Zhang, Deepika Thacker, Ting Zhou, Dazheng Zhang, Yuqing Lei, Jiajie Chen, Elizabeth Chrischilles, Dimitri A. Christakis, Soledad Fernandez, Vidu Garg, Susan Kim, Abu S. M. Mosa, Marion R. Sills, Bradley W. Taylor, David A. Williams, Qiong Wu, Christopher B. Forrest, Yong Chen the RECOVER Initiative

**Affiliations:** The Center for Health AI and Synthesis of Evidence (CHASE), University of Pennsylvania, Philadelphia, PA, USA; The Graduate Group in Applied Mathematics and Computational Science, School of Arts and Sciences, University of Pennsylvania, Philadelphia, PA, USA; Nemours Cardiac Center, Nemours Children’s Health System, Wilmington, DE, USA; Department of Biostatistics, Epidemiology, and Informatics, University of Pennsylvania Perelman School of Medicine, Philadelphia, PA, USA; Department of Epidemiology, College of Public Health, The University of Iowa, Iowa City, IA, USA; Center for Child Health, Behavior and Development, Seattle Children’s Research Institute, Seattle, WA, USA; Department of Biomedical Informatics and Center for Biostatistics, Ohio State University, Columbus, OH, USA; Heart Center and Center for Cardiovascular Research, Nationwide Children’s Hospital, Columbus, OH, USA; Department of Pediatrics, The Ohio State University, Columbus, OH, USA; Division of Pediatric Rheumatology, Benioff Children’s Hospital, University of California San Francisco, San Francisco, CA, USA; Department of Biomedical Informatics, Biostatistics and Medical Epidemiology, University of Missouri School of Medicine, Columbia, MO, USA; Department of Research, OCHIN, Inc., Portland, OR, USA; Department of Pediatrics, University of Colorado School of Medicine and Children’s Hospital Colorado, Aurora, CO, USA; Clinical and Translational Science Institute, Medical College of Wisconsin, Milwaukee, WI, USA; Department of Anesthesiology, University of Michigan, Ann Arbor, MI, USA; Applied Clinical Research Center, Department of Pediatrics, Children’s Hospital of Philadelphia, Philadelphia, PA, USA; Leonard Davis Institute of Health Economics, Philadelphia, PA, USA; Penn Medicine Center for Evidence-based Practice (CEP), Philadelphia, PA, USA; Penn Institute for Biomedical Informatics (IBI), Philadelphia, PA, USA

## Abstract

**Background:** The risk of cardiovascular outcomes in the post-acute phase of SARS-CoV-2 infection has been quantified among adults and children. This paper aimed to assess a multitude of cardiac signs, symptoms, and conditions, as well as focused on patients with and without congenital heart defects (CHDs), to provide a more comprehensive assessment of the post-acute cardiovascular outcomes among children and adolescents after COVID-19.

**Methods:** This retrospective cohort study used data from the RECOVER consortium comprising 19 US children’s hospitals and health institutions between March 2020 and September 2023. Every participant had at least a six-month follow-up after cohort entry. Absolute risks of incident post-acute COVID-19 sequelae were reported. Relative risks (RRs) were calculated by contrasting COVID-19-positive with COVID-19-negative groups using a Poisson regression model, adjusting for demographic, clinical, and healthcare utilization factors through propensity scoring stratification.

**Results:** A total of 1,213,322 individuals under 21 years old (mean[SD] age, 7.75[6.11] years; 623,806 male [51.4%]) were included. The absolute rate of any post-acute cardiovascular outcome in this study was 2.32% in COVID-19 positive and 1.38% in negative groups. Patients with CHD post-SARS-CoV-2 infection showed increased risks of any cardiovascular outcome (RR, 1.63; 95% confidence interval (CI), 1.47-1.80), including increased risks of 11 of 18 post-acute sequelae in hypertension, arrhythmias (atrial fibrillation and ventricular arrhythmias), myocarditis, other cardiac disorders (heart failure, cardiomyopathy, and cardiac arrest), thrombotic disorders (thrombophlebitis and thromboembolism), and cardiovascular-related symptoms (chest pain and palpitations). Those without CHDs also experienced heightened cardiovascular risks after SARS-CoV-2 infection (RR, 1.63; 95% CI, 1.57-1.69), covering 14 of 18 conditions in hypertension, arrhythmias (ventricular arrhythmias and premature atrial or ventricular contractions), inflammatory heart disease (pericarditis and myocarditis), other cardiac disorders (heart failure, cardiomyopathy, cardiac arrest, and cardiogenic shock), thrombotic disorders (pulmonary embolism and thromboembolism), and cardiovascular-related symptoms (chest pain, palpitations, and syncope).

**Conclusions:** Both children with and without CHDs showed increased risks for a variety of cardiovascular outcomes after SARS-CoV-2 infection, underscoring the need for targeted monitoring and management in the post-acute phase.

**Clinical Perspective section:** *What is new?:*

- We investigated the risks of 18 post-acute COVID-19 cardiovascular outcomes in the pediatric population without Multisystem Inflammatory Syndrome in Children (MIS-C) in over 1 million patients, stratified by congenital heart defects (CHD) status.
- We extended the follow-up period beyond previous pediatric studies, ensuring every participant had at least a six-month follow-up after cohort entry.
- We included a comprehensive cross-section of the US pediatric population across various healthcare settings including primary, specialty, and emergency care, as well as testing and inpatient facilities.

*What are the clinical implications?:*

- Within the post-acute phase, children and adolescents previously infected with SARS-CoV-2 are at statistically significant increased risk of incident cardiovascular outcomes, including hypertension, ventricular arrhythmias, myocarditis, heart failure, cardiomyopathy, cardiac arrest, thromboembolism, chest pain, and palpitations. These findings are consistent among patients with and without CHDs.
- Awareness of the heightened risk of cardiovascular disorders after COVID-19 can lead to a timely referral, investigations, and management of these conditions in children and adolescents.

## Introduction

Recent research has highlighted the post-acute sequelae of SARS-CoV-2 (PASC), or “long COVID”, which appears after the initial four weeks of acute infection.^1^ This condition encompasses a spectrum of symptoms affecting multiple organ systems, including cardiovascular outcomes.^2,3^ A few studies have reported increased long-term cardiovascular effects of COVID-19 in adults.^4–11^

Compared to adults, the incidence of long COVID is low in the pediatric population, including conditions such as myocarditis and general cardiorespiratory signs and symptoms.^3,12^ However, increased cardiovascular complications post-COVID-19 in the pediatric population were found in several studies, though these were limited by short follow-up periods and focused mainly on specific outcomes like myocarditis, pericarditis, and multisystem inflammatory syndrome in children associated with COVID-19 (MIS-C).^13–16^ In adults, those with pre-existing cardiovascular risk factors or established cardiovascular disease face a higher risk of severe COVID-19 and cardiovascular PASC.^17,18^ In children and adolescents, congenital heart disease (CHD) constitutes the most common congenital disorder^19^, but its effect on the risk of developing cardiovascular PASC has not been reported.

Our study utilizes data from the Researching COVID to Enhance Recovery (RECOVER) electronic health records (EHR) system to conduct a thorough evaluation of cardiovascular signs, symptoms, and conditions in children and adolescents with and without CHDs, to understand the post-acute cardiovascular impacts of COVID-19. This study has several strengths. First, this is the first and largest study to provide a comprehensive analysis of post-acute cardiovascular COVID-19 sequelae in the pediatric population. Second, the study extended the follow-up period beyond previous pediatric studies. Third, the study included a comprehensive cross-section of the US pediatric population across various healthcare settings including primary, specialty, and emergency care, as well as testing and inpatient facilities. Finally, our analysis incorporates CHDs as a key variable to enhance the precision of our findings, recognizing its prevalence as the most common congenital disorder in pediatrics.

## Methods

### Data Sources and Cohort Construction

This study is part of the National Institutes of Health (NIH) funded RECOVER Initiative (https://recovercovid.org/), which aims to learn about the long-term effects of COVID-19. This study included nineteen US children’s hospitals and health institutions. The EHR data was standardized to the PCORnet Common Data Model (CDM) and extracted from the RECOVER Database Version s10. More details are available in the Supplementary Materials.

We conducted a retrospective study from March 1, 2020, to September 1, 2023, with a cohort entry period extending from March 1, 2020, to March 6, 2023, ensuring at least a 179-day follow-up for observing post-acute COVID-19 outcomes. We included patients under 21, who had at least one healthcare visit within the baseline period, defined as 24 months to 7 days before the index date, and at least one encounter within the follow-up period, defined as 28 to 179 days after the index date.

In our study, documented SARS-CoV-2 infections were defined by positive polymerase-chain-reaction (PCR), serology, antigen tests, or diagnoses of COVID-19, or diagnoses of PASC. For the comparator group, we required all tests to be negative, no evidence or diagnosis of COVID-19 or PASC during the study period, and at least one negative COVID-19 test within the cohort entry period. The index date for COVID-19 patients was set as either the earliest date of positive tests, COVID-19 diagnoses, or 28 days before a PASC diagnosis. For COVID-19-negative patients, the index date was a randomly selected date from their negative tests to align the distribution of index dates between the two groups, controlling for time effects.

Patients diagnosed with MIS-C, Kawasaki disease, chronic kidney disease (CKD), or end-stage kidney disease (ESKD) were excluded.^20–25^ Although MIS-C is considered a PASC, it was not included in this study’s post-acute cardiovascular outcomes for two reasons: children with MIS-C can be effectively treated with minimal post-acute cardiac sequelae,^26^ and MIS-C has been and continues to be extensively studied with significantly declining incidence over the past year.^27^ **Figure 1** summarizes the participant selection for both COVID-19 positive and negative groups.

**Figure 1.**
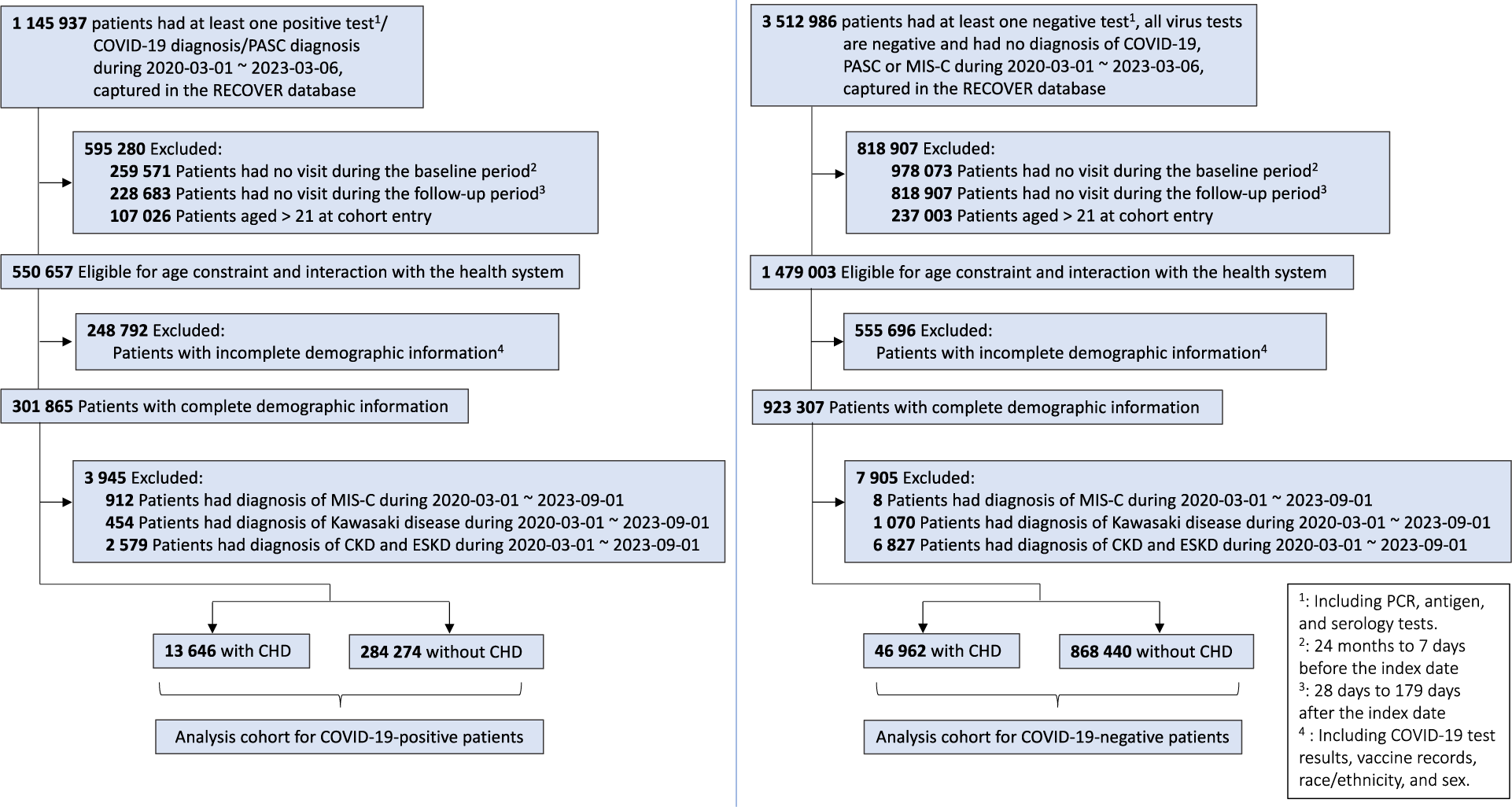
Selection of participants for both COVID-19-positive and COVID-19-negative patients, stratified by CHD status.

### Defining Cardiovascular Outcomes

We identified 18 post-acute cardiovascular outcomes for our study, including hypertension, atrial fibrillation, ventricular arrhythmias, atrial flutter, premature atrial or ventricular contractions, pericarditis, myocarditis, heart failure, cardiomyopathy, cardiac arrest, cardiogenic shock, pulmonary embolism, deep vein thrombosis, thrombophlebitis, thromboembolism, chest pain, palpitations, and syncope. These outcomes were assessed during the follow-up period in patients without a history of the specific condition during the baseline period. We used validated diagnostic codes (ICD10CM, ICD10, ICD9CM, SNOMED) confirmed by two board-certified pediatricians (DT, CF), with details of the code sets available in the Supplementary Materials.

We also grouped related cardiovascular outcomes into categories, including arrhythmias (atrial fibrillation, ventricular arrhythmias, atrial flutter, and premature atrial or ventricular contractions), inflammatory heart disease (pericarditis and myocarditis), other cardiac disorders (heart failure, cardiomyopathy, cardiac arrest, and cardiogenic shock), thrombotic disorders (pulmonary embolism, deep vein thrombosis, thrombophlebitis, and thromboembolism), cardiovascular-related symptoms (chest pain, palpitations, and syncope), and any cardiovascular outcome (any incident cardiovascular condition studied).

### Covariates

We examined a comprehensive set of patient characteristics as measured confounders collected before cohort entry, to be adjusted through propensity score stratification^28^ to balance the comparison groups. These included demographic factors, including age at index date, gender (female, male), and race/ethnicity (Non-Hispanic White (NHW), Non-Hispanic Black (NHB), Hispanic, Asian American/Pacific Islander (AAPI), Multiple, Other/Unknown); clinical factors, including obesity status, a chronic condition indicator defined by the Pediatric Medical Complexity Algorithm^29^ (PMCA, no chronic condition, non-complex chronic condition, complex chronic condition), and a list of pre-existing chronic conditions^3,30^; health care utilization factors collected 24 months to 7 days before index date, including the number of inpatient visits, outpatient visits, emergency department (ED) visits, unique medications, and negative COVID-19 tests (0, 1, 2, ≥3); vaccine information, including dosage of COVID-19 vaccine before index date (0, 1, ≥2) and interval since the last COVID-19 immunization (no vaccine, <4 months, ≥ 4 months); year-month of cohort entry (from March 2020 to March 2023); indicators from the 19 data-contributing sites.

### Statistical Analysis

We calculated the incidence of post-acute cardiovascular outcomes in COVID-19-positive and negative cohorts, stratified by CHD status. For each outcome, incidence rates were calculated by dividing new cases during the follow-up period by the total number of patients, excluding those with the specific outcome at baseline.

We presented distributions of preference scores—a transformation of propensity scores accounting for prevalence differences between populations—to assess empirical equipoise^31^. Empirical equipoise is achieved for COVID-19 positive and negative patients when the majority of individuals in both groups have preference scores ranging from 0.3 to 0.7.^28,31^ To mitigate the effects of confounding, we used propensity score stratification^28^ to adjust for a large number of measured confounders collected before index date. After stratification, we assessed the standardized mean difference (SMD) of each covariate between COVID-19 positive and negative patients, with an SMD of 0.1 or less indicating acceptable balance.^28,32^

We used the stratified Poisson regression model to estimate the relative risk (RR) for each outcome between comparison groups for patients with and without CHD. RR is a collapsible measure, meaning the measure of association conditional on some factors remains consistent with the marginal measure collapsed over strata, which is crucial for accurate interpretation in clinical research.^33,34^

We also conducted subgroup analyses by age (0-4, 5-11, 12-20 years), race/ethnicity (NHW, NHB, Hispanic), gender (male and female), obesity status (obese and non-obese), severity of acute COVID-19^35^ (“non-severe” including asymptomatic and mild, “severe” including moderate and severe), and estimated time frames corresponding to dominant virus variants (pre-Delta, Delta, Omicron). Specifically, the pre-Delta variant spanned March 1, 2021, to June 30, 2021; Delta from July 1, 2021, to December 31, 2021; and Omicron from January 1, 2022, to March 6, 2023, with a minimum 179-day follow-up to observe long COVID outcomes.^36^

### Sensitivity Analysis

We conducted extensive sensitivity analyses to examine the robustness of our findings. Negative control outcome experiments^28,37,38^ were performed to calibrate the residual study bias from unmeasured confounders and systematic sources, in which the null hypothesis of no effect was believed to be true utilizing a list of 36 negative control outcomes determined by two board-certified pediatricians (DT, CF). The empirical null distribution and calibrated risks were reported in Supplementary Materials **Section S3**. Additional analyses were performed excluding patients included solely based on a PASC diagnosis (**Section S4**) and those without any cardiovascular outcomes within the baseline period (**Section S5**). Given limited SARS-CoV-2 testing availability during the first wave of COVID-19 (March to May 2020), we conducted analyses excluding patients whose index dates fell within this period (**Section S6**). Furthermore, we excluded data from site L where the population is truncated at the ambulatory level (**Section S7**). All analyses were performed using R version 4.0.2.

## Results

### Cohort Identification

A total of 297,920 children and adolescents with COVID-19 (24.6%; 13,646 with CHDs, 284,274 without) and 915,402 without COVID-19 (75.4%; 46,962 with CHDs, 868,440 without) in the RECOVER Database were included. Among these patients, 623,806 (51.4%) were male with a mean (SD) age of 7.75 (6.11) years. Baseline characteristics by COVID-19 and CHD status are detailed in **Table 1**.

**Table 1.** Baseline characteristics of patients, stratified by COVID-19 status and CHD status.

|  | *COVID-19-positive* |  |  | *COVID-19-negative* |  |  |
| --- | --- | --- | --- | --- | --- | --- |
|  | **CHD**<br>**(N=13,646)** | **Non-CHD**<br>**(N=284,274)** | **Overall**<br>**(N=297,920)** | **CHD**<br>**(N=46,962)** | **Non-CHD**<br>**(N=868,440)** | **Overall**<br>**(N=915,402)** |
| **Age** |  |  |  |  |  |  |
| Median [Q1, Q3] | 3.0 [1.0, 8.0] | 8.0 [2.0, 14.0] | 8.0 [2.0, 14.0] | 3.0 [1.0, 8.0] | 7.0 [2.0, 13.0] | 6.0 [2.0, 13.0] |
| **Year of Age, no. (%)** |  |  |  |  |  |  |
| 0 | 3,126 (22.9) | 35,297 (12.4) | 38,423 (12.9) | 11,632 (24.8) | 90,104 (10.4) | 101,736 (11.1) |
| 1 | 1,879 (13.8) | 22,792 (8.0) | 24,671 (8.3) | 5,486 (11.7) | 80,038 (9.2) | 85,524 (9.3) |
| 2 | 1,214 (8.9) | 16,011 (5.6) | 17,225 (5.8) | 3,765 (8.0) | 60,131 (6.9) | 63,896 (7.0) |
| 3 | 1,007 (7.4) | 13,679 (4.8) | 14,686 (4.9) | 3,547 (7.6) | 54,274 (6.2) | 57,821 (6.3) |
| 4 | 810 (5.9) | 13,439 (4.7) | 14,249 (4.8) | 3,111 (6.6) | 52,261 (6.0) | 55,372 (6.0) |
| 5 | 725 (5.3) | 12,248 (4.3) | 12,973 (4.4) | 2,856 (6.1) | 50,846 (5.9) | 53,702 (5.9) |
| 6 | 588 (4.3) | 11,603 (4.1) | 12,191 (4.1) | 2,292 (4.9) | 44,811 (5.2) | 47,103 (5.1) |
| 7 | 491 (3.6) | 11,270 (4.0) | 11,761 (3.9) | 1,839 (3.9) | 39,620 (4.6) | 41,459 (4.5) |
| 8 | 429 (3.1) | 11,105 (3.9) | 11,534 (3.9) | 1,513 (3.2) | 35,983 (4.1) | 37,496 (4.1) |
| 9 | 410 (3.0) | 11,229 (4.0) | 11,639 (3.9) | 1,358 (2.9) | 33,960 (3.9) | 35,318 (3.9) |
| 10 | 370 (2.7) | 11,820 (4.2) | 12,190 (4.1) | 1,213 (2.6) | 32,537 (3.7) | 33,750 (3.7) |
| 11 | 374 (2.7) | 12,532 (4.4) | 12,906 (4.3) | 1,174 (2.5) | 32,381 (3.7) | 33,555 (3.7) |
| 12 | 282 (2.1) | 10,969 (3.9) | 11,251 (3.8) | 989 (2.1) | 31,364 (3.6) | 32,353 (3.5) |
| 13 | 276 (2.0) | 11,583 (4.1) | 11,859 (4.0) | 993 (2.1) | 32,054 (3.7) | 33,047 (3.6) |
| 14 | 275 (2.0) | 12,273 (4.3) | 12,548 (4.2) | 974 (2.1) | 33,222 (3.8) | 34,196 (3.7) |
| 15 | 277 (2.0) | 13,051 (4.6) | 13,328 (4.5) | 962 (2.0) | 33,911 (3.9) | 34,873 (3.8) |
| 16 | 278 (2.0) | 13,268 (4.7) | 13,546 (4.5) | 896 (1.9) | 34,232 (3.9) | 35,128 (3.8) |
| 17 | 275 (2.0) | 13,181 (4.6) | 13,456 (4.5) | 825 (1.8) | 33,998 (3.9) | 34,823 (3.8) |
| 18 | 225 (1.6) | 9,695 (3.4) | 9,920 (3.3) | 615 (1.3) | 24,962 (2.9) | 25,577 (2.8) |
| 19 | 160 (1.2) | 8,725 (3.1) | 8,885 (3.0) | 480 (1.0) | 19,826 (2.3) | 20,306 (2.2) |
| 20 | 175 (1.3) | 8,504 (3.0) | 8,679 (2.9) | 442 (0.9) | 17,925 (2.1) | 18,367 (2.0) |
| **Gender, no. (%)** |  |  |  |  |  |  |
| Female | 6,333 (46.4) | 141,868 (49.9) | 148,201 (49.7) | 22,066 (47.0) | 419,249 (48.3) | 441,315 (48.2) |
| Male | 7,313 (53.6) | 142,406 (50.1) | 149,719 (50.3) | 24,896 (53.0) | 449,191 (51.7) | 474,087 (51.8) |
| **Race/Ethnicity, no. (%)** |  |  |  |  |  |  |
| NHW | 6,247 (45.8) | 118,054 (41.5) | 124,301 (41.7) | 23,067 (49.1) | 395,378 (45.5) | 418,445 (45.7) |
| NHB | 2,307 (16.9) | 51,139 (18.0) | 53,446 (17.9) | 7,294 (15.5) | 147,576 (17.0) | 154,870 (16.9) |
| Hispanic | 3,197 (23.4) | 70,834 (24.9) | 74,031 (24.8) | 9,662 (20.6) | 187,025 (21.5) | 196,687 (21.5) |
| AAPI | 586 (4.3) | 13,573 (4.8) | 14,159 (4.8) | 2,124 (4.5) | 41,659 (4.8) | 43,783 (4.8) |
| Multiple | 400 (2.9) | 7,493 (2.6) | 7,893 (2.6) | 1,486 (3.2) | 26,357 (3.0) | 27,843 (3.0) |
| Other/Unknown | 909 (6.7) | 23,181 (8.2) | 24,090 (8.1) | 3,329 (7.1) | 70,445 (8.1) | 73,774 (8.1) |

| Health System, no. (%) |  |  |  |  |  |  |
| --- | --- | --- | --- | --- | --- | --- |
| A | 1,212 (8.9) | 26,724 (9.4) | 27,936 (9.4) | 4,259 (9.1) | 86,746 (10.0) | 91,005 (9.9) |
| B | 1,730 (12.7) | 52,598 (18.5) | 54,328 (18.2) | 6,099 (13.0) | 134,278 (15.5) | 140,377 (15.3) |
| C | 1,251 (9.2) | 13,977 (4.9) | 15,228 (5.1) | 4,890 (10.4) | 61,282 (7.1) | 66,172 (7.2) |
| D | 99 (0.7) | 5,222 (1.8) | 5,321 (1.8) | 255 (0.5) | 10,480 (1.2) | 10,735 (1.2) |
| E | 838 (6.1) | 7,171 (2.5) | 8,009 (2.7) | 3,234 (6.9) | 35,653 (4.1) | 38,887 (4.2) |
| F | 702 (5.1) | 12,351 (4.3) | 13,053 (4.4) | 2,806 (6.0) | 47,709 (5.5) | 50,515 (5.5) |
| G | 431 (3.2) | 6,942 (2.4) | 7,373 (2.5) | 872 (1.9) | 16,838 (1.9) | 17,710 (1.9) |
| H | 606 (4.4) | 9,178 (3.2) | 9,784 (3.3) | 1,816 (3.9) | 28,694 (3.3) | 30,510 (3.3) |
| I | 344 (2.5) | 5,121 (1.8) | 5,465 (1.8) | 1,041 (2.2) | 13,666 (1.6) | 14,707 (1.6) |
| J | 1,165 (8.5) | 22,316 (7.9) | 23,481 (7.9) | 4,845 (10.3) | 102,912 (11.9) | 107,757 (11.8) |
| K | 1,154 (8.5) | 23,754 (8.4) | 24,908 (8.4) | 4,048 (8.6) | 85,730 (9.9) | 89,778 (9.8) |
| L | 593 (4.3) | 55,135 (19.4) | 55,728 (18.7) | 1,180 (2.5) | 116,396 (13.4) | 117,576 (12.8) |
| M | 539 (3.9) | 8,408 (3.0) | 8,947 (3.0) | 1,259 (2.7) | 14,638 (1.7) | 15,897 (1.7) |
| O | 8 (0.1) | 673 (0.2) | 681 (0.2) | 27 (0.1) | 1,693 (0.2) | 1,720 (0.2) |
| P | 593 (4.3) | 4,143 (1.5) | 4,736 (1.6) | 2,359 (5.0) | 24,422 (2.8) | 26,781 (2.9) |
| Q | 502 (3.7) | 7,947 (2.8) | 8,449 (2.8) | 2,069 (4.4) | 24,457 (2.8) | 26,526 (2.9) |
| R | 10 (0.1) | 402 (0.1) | 412 (0.1) | 7 (0.0) | 317 (0.0) | 324 (0.0) |
| S | 798 (5.8) | 10,723 (3.8) | 11,521 (3.9) | 2,638 (5.6) | 34,231 (3.9) | 36,869 (4.0) |
| T | 1,071 (7.8) | 11,489 (4.0) | 12,560 (4.2) | 3,258 (6.9) | 28,298 (3.3) | 31,556 (3.4) |
| Entry time, no. (%) |  |  |  |  |  |  |
| 03/2020 - 05/2020 | 167 (1.2) | 2,708 (1.0) | 2,875 (1.0) | 1,545 (3.3) | 15,812 (1.8) | 17,357 (1.9) |
| 06/2020 - 08/2020 | 308 (2.3) | 9,681 (3.4) | 9,989 (3.4) | 4,028 (8.6) | 56,525 (6.5) | 60,553 (6.6) |
| 09/2020 - 11/2020 | 588 (4.3) | 16,600 (5.8) | 17,188 (5.8) | 4,305 (9.2) | 78,031 (9.0) | 82,336 (9.0) |
| 12/2020 - 02/2021 | 1,025 (7.5) | 28,124 (9.9) | 29,149 (9.8) | 3,761 (8.0) | 75,910 (8.7) | 79,671 (8.7) |
| 03/2021 - 05/2021 | 729 (5.3) | 16,304 (5.7) | 17,033 (5.7) | 4,316 (9.2) | 79,840 (9.2) | 84,156 (9.2) |
| 06/2021 - 08/2021 | 726 (5.3) | 16,924 (6.0) | 17,650 (5.9) | 4,466 (9.5) | 78,411 (9.0) | 82,877 (9.1) |
| 09/2021 - 11/2021 | 1,140 (8.4) | 26,733 (9.4) | 27,873 (9.4) | 5,259 (11.2) | 120,200 (13.8) | 125,459 (13.7) |
| 12/2021 - 02/2022 | 3,736 (27.4) | 82,586 (29.1) | 86,322 (29.0) | 4,243 (9.0) | 89,917 (10.4) | 94,160 (10.3) |
| 03/2022 - 05/2022 | 1,188 (8.7) | 20,014 (7.0) | 21,202 (7.1) | 3,770 (8.0) | 67,148 (7.7) | 70,918 (7.7) |
| 06/2022 - 08/2022 | 1,889 (13.8) | 32,369 (11.4) | 34,258 (11.5) | 3,172 (6.8) | 52,367 (6.0) | 55,539 (6.1) |
| 09/2022 - 11/2022 | 956 (7.0) | 15,140 (5.3) | 16,096 (5.4) | 4,291 (9.1) | 83,680 (9.6) | 87,971 (9.6) |
| 12/2022 - 03/2023 | 1,194 (8.7) | 17,091 (6.0) | 18,285 (6.1) | 3,806 (8.1) | 70,599 (8.1) | 74,405 (8.1) |
| Obesity, no. (%) |  |  |  |  |  |  |
| No | 8,861 (64.9) | 149,365 (52.5) | 158,226 (53.1) | 31,985 (68.1) | 495,600 (57.1) | 527,585 (57.6) |
| Yes | 4,425 (32.4) | 110,883 (39.0) | 115,308 (38.7) | 13,057 (27.8) | 288,267 (33.2) | 301,324 (32.9) |
| Unknown | 360 (2.6) | 24,026 (8.5) | 24,386 (8.2) | 1,920 (4.1) | 84,573 (9.7) | 86,493 (9.4) |
| **PMCA, no. (%)** |  |  |  |  |  |  |
| 0 | 3,172 (23.2) | 202,897 (71.4) | 206,069 (69.2) | 12,383 (26.4) | 619,127 (71.3) | 631,510 (69.0) |
| 1 | 2,544 (18.6) | 48,078 (16.9) | 50,622 (17.0) | 8,759 (18.7) | 146,594 (16.9) | 155,353 (17.0) |
| 2 | 7,930 (58.1) | 33,299 (11.7) | 41,229 (13.8) | 25,820 (55.0) | 102,719 (11.8) | 128,539 (14.0) |
| **Number of negative COVID-19 tests before index date, no. (%)** |  |  |  |  |  |  |
| 0 | 5,633 (41.3) | 169,814 (59.7) | 175,447 (58.9) | 29,732 (63.3) | 649,004 (74.7) | 678,736 (74.1) |
| 1 | 2,855 (20.9) | 60,909 (21.4) | 63,764 (21.4) | 8,764 (18.7) | 141,107 (16.2) | 149,871 (16.4) |
| 2 | 1,605 (11.8) | 25,685 (9.0) | 27,290 (9.2) | 3,637 (7.7) | 44,266 (5.1) | 47,903 (5.2) |
| ≥3 | 3,553 (26.0) | 27,866 (9.8) | 31,419 (10.5) | 4,829 (10.3) | 34,063 (3.9) | 38,892 (4.2) |
| **Dosage of COVID-19 vaccine, no. (%)** |  |  |  |  |  |  |
| 0 | 9,995 (73.2) | 175,674 (61.8) | 185,669 (62.3) | 34,796 (74.1) | 566,168 (65.2) | 600,964 (65.7) |
| 1 | 773 (5.7) | 17,218 (6.1) | 17,991 (6.0) | 2,336 (5.0) | 46,143 (5.3) | 48,479 (5.3) |
| ≥2 | 2,878 (21.1) | 91,382 (32.1) | 94,260 (31.6) | 9,830 (20.9) | 256,129 (29.5) | 265,959 (29.1) |
| **Number of unique medications, no. (%)** |  |  |  |  |  |  |
| 0 | 1,458 (10.7) | 61,432 (21.6) | 62,890 (21.1) | 7,221 (15.4) | 224,089 (25.8) | 231,310 (25.3) |
| 1 | 768 (5.6) | 31,344 (11.0) | 32,112 (10.8) | 3,397 (7.2) | 100,046 (11.5) | 103,443 (11.3) |
| 2 | 646 (4.7) | 26,711 (9.4) | 27,357 (9.2) | 2,580 (5.5) | 80,440 (9.3) | 83,020 (9.1) |
| ≥3 | 10,774 (79.0) | 164,787 (58.0) | 175,561 (58.9) | 33,764 (71.9) | 463,865 (53.4) | 497,629 (54.4) |

### Incidence of Post-acute Cardiovascular Events

**Table 2** presents the incidence of 18 individual and 6 composite post-acute cardiovascular outcomes for COVID-19-positive compared to COVID-19-negative patients, stratified by CHD status. The data indicate that COVID-19-positive patients generally have higher incidences in both CHD and non-CHD groups. For example, the absolute risk of heart failure was 1.61% in COVID-19-positive patients with CHD, compared to 1.18% in their COVID-19-negative counterparts. Similarly, chest pain incidence was 1.23% in COVID-19-positive patients without CHD, versus 0.61% in COVID-19-negative patients. Overall, the CHD group showed higher absolute risks of any post-acute cardiovascular outcomes than the non-CHD group, with 5.57% for positive and 4.00% for negative patients with CHD, compared to 2.19% and 1.26% respectively in those without CHD.

**Table 2.** Absolute risks (in %, incident/total patients) of individual and composite cardiovascular outcomes, stratified by COVID-19 status and CHD status.

|  | *COVID-19-positive* |  |  | *COVID-19-negative* |  |  |
| --- | --- | --- | --- | --- | --- | --- |
|  | **CHD**<br>**(N=13,646)** | **Non-CHD**<br>**(N=284,274)** | **Overall**<br>**(N=297,920)** | **CHD**<br>**(N=46,962)** | **Non-CHD**<br>**(N=868,440)** | **Overall**<br>**(N=915,402)** |
| **Any cardiovascular outcome** | 5.566 (601/10797) | 2.19 (5895/269210) | 2.32 (6496/280007) | 3.998 (1529/38248) | 1.261 (10481/831098) | 1.381 (12010/869346) |
| Hypertension | 1.531 (195/12736) | 0.333 (936/281427) | 0.384 (1131/294163) | 1.08 (481/44548) | 0.209 (1801/861469) | 0.252 (2282/906017) |
| **Any arrhythmias** | 0.967 (126/13029) | 0.073 (208/283655) | 0.113 (334/296684) | 0.79 (355/44955) | 0.053 (461/866011) | 0.09 (816/910966) |
| Atrial fibrillation | 0.081 (11/13619) | 0.006 (16/284233) | 0.009 (27/297852) | 0.038 (18/46896) | 0.005 (46/868329) | 0.007 (64/915225) |
| Ventricular arrhythmias | 0.863 (113/13089) | 0.06 (171/283757) | 0.096 (284/296846) | 0.695 (314/45159) | 0.044 (378/866270) | 0.076 (692/911429) |
| Atrial flutter | 0.059 (8/13577) | 0.002 (7/284243) | 0.005 (15/297820) | 0.092 (43/46754) | 0.002 (17/868365) | 0.007 (60/915119) |
| Premature atrial or ventricular contractions | 0.059 (8/13619) | 0.009 (26/284206) | 0.011 (34/297825) | 0.058 (27/46862) | 0.006 (52/868245) | 0.009 (79/915107) |
| **Any inflammatory heart disease** | 0.125 (17/13603) | 0.044 (126/284132) | 0.048 (143/297735) | 0.094 (44/46828) | 0.015 (134/868141) | 0.019 (178/914969) |
| Pericarditis | 0.081 (11/13628) | 0.014 (39/284198) | 0.017 (50/297826) | 0.077 (36/46886) | 0.007 (64/868296) | 0.011 (100/915182) |
| Myocarditis | 0.059 (8/13620) | 0.034 (97/284185) | 0.035 (105/297805) | 0.019 (9/46902) | 0.01 (83/868259) | 0.01 (92/915161) |
| **Any other cardiac disorders** | 2.236 (281/12565) | 0.079 (225/283615) | 0.171 (506/296180) | 1.632 (710/43513) | 0.055 (474/866289) | 0.13 (1184/909802) |
| Heart failure | 1.609 (208/12930) | 0.02 (57/284114) | 0.089 (265/297044) | 1.179 (527/44688) | 0.012 (106/868010) | 0.069 (633/912698) |
| Cardiomyopathy | 0.578 (77/13331) | 0.042 (118/283824) | 0.066 (195/297155) | 0.361 (166/46046) | 0.028 (245/867069) | 0.045 (411/913115) |
| Cardiac arrest | 0.483 (65/13461) | 0.032 (91/284122) | 0.052 (156/297583) | 0.356 (165/46303) | 0.022 (194/867807) | 0.039 (359/914110) |
| Cardiogenic shock | 0.155 (21/13526) | 0.006 (17/284233) | 0.013 (38/297759) | 0.12 (56/46554) | 0.002 (18/868332) | 0.008 (74/914886) |
| **Any thrombotic disorders** | 0.913 (119/13029) | 0.102 (288/283427) | 0.137 (407/296456) | 0.737 (332/45039) | 0.084 (728/865935) | 0.116 (1060/910974) |
| Pulmonary embolism | 0.103 (14/13618) | 0.022 (62/284178) | 0.026 (76/297796) | 0.047 (22/46872) | 0.013 (110/868175) | 0.014 (132/915047) |
| Deep vein thrombosis | 0.096 (13/13585) | 0.011 (31/284188) | 0.015 (44/297773) | 0.062 (29/46848) | 0.01 (86/868264) | 0.013 (115/915112) |
| Thrombophlebitis | 0.096 (13/13591) | 0.018 (51/284141) | 0.021 (64/297732) | 0.036 (17/46817) | 0.017 (151/867965) | 0.018 (168/914782) |
| Thromboembolism | 0.896 (117/13064) | 0.091 (258/283513) | 0.126 (375/296577) | 0.72 (325/45113) | 0.074 (639/866229) | 0.106 (964/911342) |
| **Cardiovascular-related symptoms** | 2.002 (259/12938) | 1.748 (4772/272941) | 1.76 (5031/285879) | 1.028 (463/45035) | 0.959 (8071/842021) | 0.962 (8534/887056) |
| Chest pain | 1.345 (177/13160) | 1.229 (3401/276688) | 1.234 (3578/289848) | 0.637 (292/45810) | 0.611 (5202/851857) | 0.612 (5494/897667) |
| Palpitations | 0.564 (76/13474) | 0.378 (1067/282181) | 0.387 (1143/295655) | 0.274 (127/46402) | 0.181 (1566/863436) | 0.186 (1693/909838) |
| Syncope | 0.371 (50/13482) | 0.42 (1181/281122) | 0.418 (1231/294604) | 0.271 (126/46454) | 0.298 (2563/860375) | 0.297 (2689/906829) |

### Adjusted Relative Risk of post-acute Cardiovascular Outcomes

The COVID-19 positive and negative patients achieved empirical equipoise (**Figure S2**), with well-balanced characteristics after propensity score stratification (**Figure S3**). **Figure 2** highlights that post-acute cardiovascular outcomes are higher in both CHD and non-CHD groups among patients with COVID-19 compared to negative ones.

**Figure 2.**
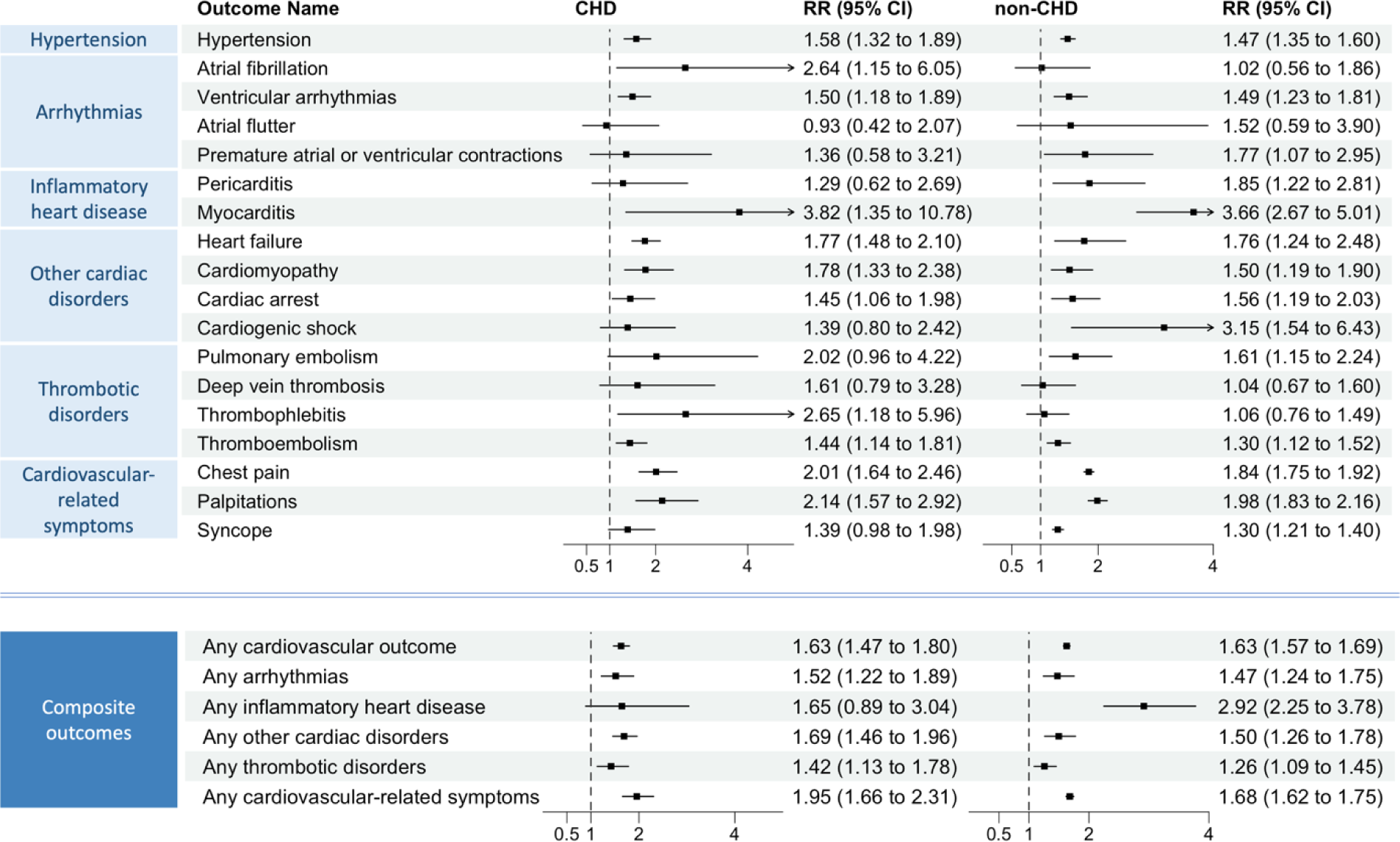
Relative Risk (RR) of incident post-acute COVID-19 cardiovascular outcomes compared with the COVID-19-negative cohort, stratified by CHD status. Composite outcomes consisted of arrhythmias (atrial fibrillation, ventricular arrhythmias, atrial flutter, and premature atrial or ventricular contractions), inflammatory heart disease (pericarditis and myocarditis), other cardiac disorders (heart failure, cardiomyopathy, cardiac arrest, and cardiogenic shock), thrombotic disorders (pulmonary embolism, deep vein thrombosis, thrombophlebitis, and thromboembolism), cardiovascular-related symptoms (chest pain, palpitations, and syncope), and any cardiovascular outcome (incident occurrence of any cardiovascular outcome studied).

Composite outcomes demonstrated increased risks in both groups. For CHD patients, there were increases across any cardiovascular outcome (RR, 1.63; 95% CI, 1.47-1.80), arrhythmias (RR, 1.52; 95% CI, 1.22-1.89), other cardiac disorders (RR, 1.69; 95% CI, 1.46-1.96), thrombotic disorders (RR, 1.42; 95% CI, 1.13-1.78), and cardiovascular-related symptoms (RR, 1.95; 95% CI, 1.66-2.31). For non-CHD patients, increases were noted in any cardiovascular outcome (RR, 1.63; 95% CI, 1.57-1.69), arrhythmias (RR, 1.47; 95% CI, 1.24-1.75), inflammatory heart disease (RR, 2.92; 95% CI, 2.25-3.78), other cardiac disorders (RR, 1.50; 95% CI, 1.26-1.78), thrombotic disorders (RR, 1.26; 95% CI, 1.09-1.45), and cardiovascular-related symptoms (RR, 1.68; 95% CI, 1.62-1.75).

Individual outcomes also showed increased risks. CHD patients experienced higher risks in hypertension (RR, 1.58; 95% CI, 1.32-1.89), atrial fibrillation (RR, 2.64; 95% CI, 1.15-6.05), ventricular arrhythmias (RR, 1.50; 95% CI, 1.18-1.89), myocarditis (RR, 3.82; 95% CI, 1.35-10.78), heart failure (RR, 1.77; 95% CI, 1.48-2.10), cardiomyopathy (RR, 1.78, 95% CI, 1.33-2.38), cardiac arrest (RR, 1.45; 95% CI, 1.06-1.98), thrombophlebitis (RR, 2.65; 95% CI, 1.18-5.96), thromboembolism (RR, 1.44; 95% CI, 1.14-1.81), chest pain (RR, 2.01; 95% CI, 1.64-2.46), and palpitations (RR, 2.14; 95% CI, 1.57-2.92). Non-CHD patients saw increased risks in hypertension (RR, 1.47; 95% CI, 1.35-1.60), ventricular arrhythmias (RR, 1.49; 95% CI, 1.23-1.81), premature atrial or ventricular contractions (RR, 1.77; 95% CI 1.07-2.95), pericarditis (RR, 1.85; 95% CI, 1.22-2.81), myocarditis (RR, 3.66; 95% CI, 2.67-5.01), heart failure (RR, 1.76; 95% CI, 1.24-2.48), cardiomyopathy (RR, 1.50; 95% CI, 1.19-1.90), cardiac arrest (RR, 1.56; 95% CI, 1.19-2.03), cardiogenic shock (RR, 3.15; 95% CI, 1.54-6.43), pulmonary embolism (RR, 1.61; 95% CI, 1.15-2.24), thromboembolism (RR, 1.30; 95% CI, 1.12-1.52), chest pain (RR, 1.84; 95% CI, 1.75-1.92), palpitations (RR, 1.98; 95% CI, 1.83-2.16), and syncope (RR, 1.30; 95% CI, 1.21-1.40).

Figure 3 demonstrated that the risks of post-acute cardiovascular composite outcomes were evident across age groups, race/ethnicity, gender, obesity status, severity of acute COVID-19, and dominant virus variants.

**Figure 3.**
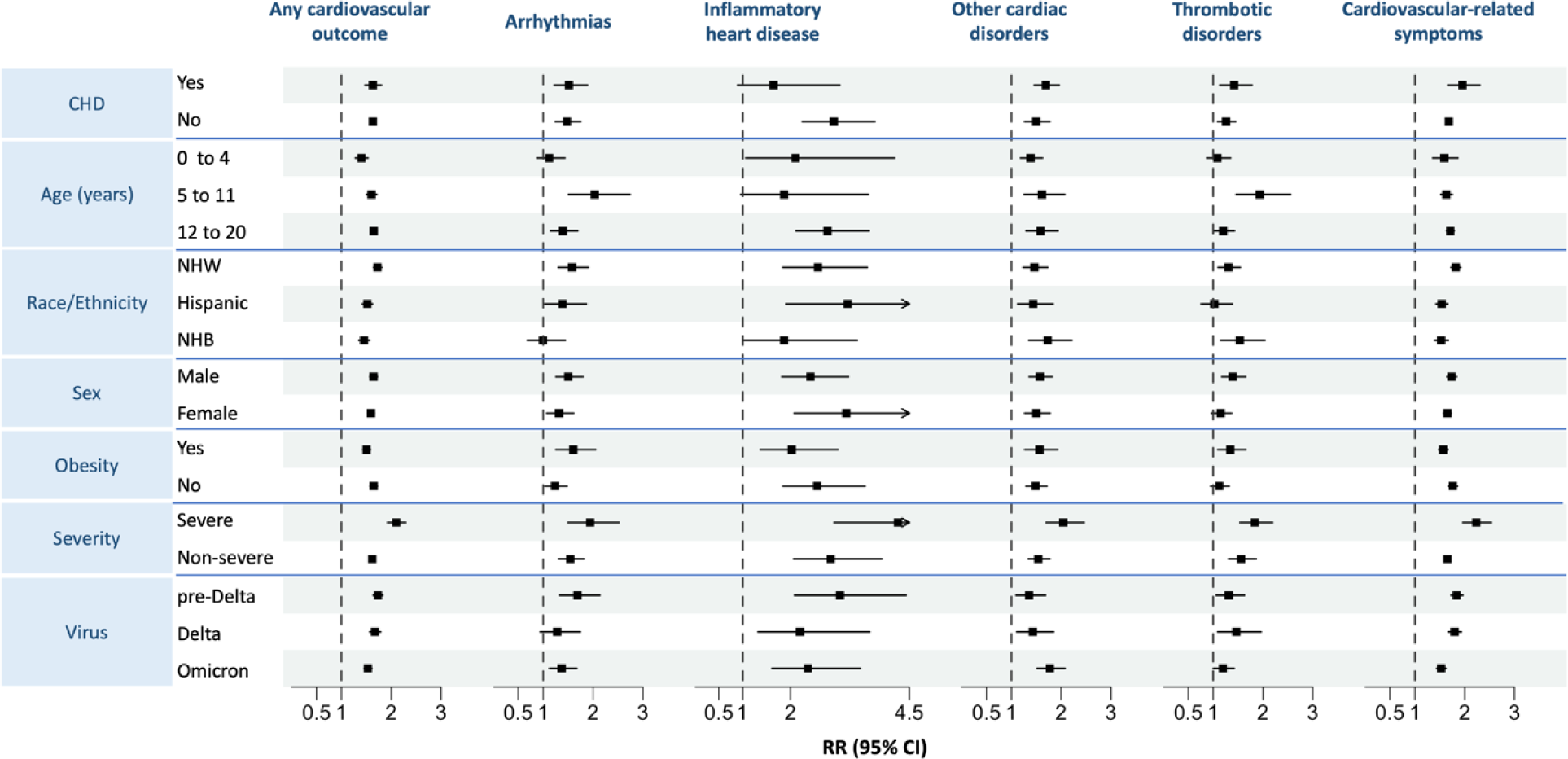
Subgroup analyses of Relative Risk (RR) of incident post-acute COVID-19 composite cardiovascular outcomes compared with the COVID-19-negative cohort, according to CHD status, age, race/ethnicity, sex, obesity, severity during the acute phase of COVID-19, and the dominant variant.

### Sensitivity Analysis

Negative control experiments (**Section S3**) indicated a slight systematic error, as shown by a minor shift in point estimates with wider CIs. Analyses excluding patients solely based on PASC (**Section S4**), those without prior cardiovascular history within the baseline period (**Section S5**), those from the first COVID-19 wave (March to May 2020, **Section S6**), and those at site L (**Section S7**), all gave similar results as in the primary analyses. Adolescents over 12 and children aged 5 to 11 displayed higher risks of any cardiovascular outcomes compared to children under 5 (**Section S8**). NHW patients exhibited the highest risks of any cardiovascular outcome, followed by Hispanic and NHB groups (**Section S9**). Gender differences were noted: females showed higher risks for hypertension, deep vein thrombosis, and cardiac arrest, whereas males were more prone to premature atrial or ventricular contractions, pulmonary embolism, and thrombophlebitis (**Section S10**). Obesity was associated with increased risks of ventricular arrhythmias, premature atrial or ventricular contractions, cardiogenic shock, pulmonary embolism, and thrombophlebitis, but lower risks of cardiovascular-related symptoms (**Section S11**). Patients with severe COVID-19 during the acute phase had higher risks across all composite cardiovascular outcomes compared to the non-severe group (**Section S12**). The pre-Delta period saw the highest risk of any cardiovascular outcome, with consistent risks observed across all dominant virus variants (**Section S13**).

## Discussion

Our study involved 297,920 children and adolescents with COVID-19 and 915,402 without COVID-19. We found that those infected with SARS-CoV-2 exhibited increased risks for a range of post-acute cardiovascular outcomes, with RR between 1.26 and 2.92. These outcomes included hypertension, ventricular arrhythmias, myocarditis, heart failure, cardiomyopathy, cardiogenic shock, thromboembolism, chest pain, and palpitations, compared to non-infected controls. These findings applied to patients both with and without CHDs, although children with CHD showed a notably higher risk of atrial fibrillation. Especially noteworthy was the increased risk of myocarditis in both groups consistent with prior studies showing an increased occurrence of myocarditis in the months following the diagnosis of COVID-19, across different ages and genders ^3,39–43^. Risks were consistently observed regardless of age, gender, race/ethnicity, obesity status, severity of acute COVID-19, or virus variant. Even children and adolescents without a history of any cardiovascular outcomes before SARS-CoV-2 infection showed increased risks, suggesting a broad potential impact on those previously considered at low risk of cardiovascular disease. Our results were robust through extensive sensitivity analyses and negative control experiments.

Our study has several strengths. First, we used the RECOVER EHR database to build a large cohort with and without COVID exposure, providing longitudinal follow-up throughout the post-acute period, which extended past findings in both populations and outcomes definitions. Second, propensity score stratification, accommodating hundreds of covariates while balancing the covariates in the two comparative groups, improved confounder adjustment over traditional linear regression, and reduced non-linear confounder effects^44^. Third, all analyses were stratified by CHD status, a unique consideration in the pediatric setting. We also employed RR as our comparative measure, important for its collapsibility^34^ and accurate interpretation in clinical research^33^. Lastly, we conducted negative control experiments^37,38^ to address systematic bias and control unmeasured confounders.

The COVID-19 pandemic prompted the widespread use of social distancing, masking, subsequently vaccines, and medications. As acute cases declined, attention shifted towards long-term sequelae like cardiovascular PASC, particularly significant in pediatric patients, including competitive athletes with a history of COVID-19 and any subsequent cardiac complications^45,46^. The substantial risks of post-acute cardiovascular effects of SARS-CoV-2 in pediatrics call for dedicated healthcare resource allocation and long-term monitoring focused on cardiovascular health.

## Limitations

This study has several limitations. First, selecting high-quality COVID-19 controls is challenging due to potential misclassification^47^; to mitigate this, we incorporated a combination of PCR, antigen, and serology tests, along with diagnosis codes for COVID-19 and long COVID, to define our COVID-19 control group more accurately. On the other hand, although historical or pre-pandemic controls have been suggested, this introduces distinct difficulties for pediatric research due to the dynamic nature of rapid physical growth and physiological, cognitive, emotional, and social development in children and adolescents, which may not align with pre-pandemic benchmarks and could potentially lead to misleading results. Therefore, we used contemporary controls in our primary analyses for a valid comparison. In addition, the increased use of at-home rapid antigen tests later in the pandemic may have resulted in underreported testing frequencies in EHRs. Nonetheless, our sensitivity analyses across different pandemic phases were consistent, showing resilience to the influence of viral variants and mitigating concerns about data capture completeness.

Second, observational studies inherently face the challenge of confounding bias. To mitigate this, we used a propensity-score-based stratification approach and included a large number of potential confounders. We also incorporated the negative control experiments^37,38^ in our sensitivity analyses to adjust for unmeasured confounders. However, EHR data may still suffer from misclassification biases, such as incorrect documentation of SARS-CoV-2 infection status or incomplete follow-up data. Some efforts to lessen these biases have been made.^37,38,48,49^

Third, the frequency of hospital visits^50^ by COVID-19-positive patients might bias the observed post-acute cardiovascular outcomes upward, as these patients may visit hospitals more frequently. We tried to address this by including healthcare utilization factors such as hospital visits and tests as confounders to balance the comparison groups.

Fourth, our analysis did not account for reinfection or COVID-19 vaccinations during the study period, which could affect outcomes like myocarditis or pericarditis.^13,51–53^ We included the dosage of COVID-19 vaccines and the interval since the last immunization as confounders to partially adjust for these variables, but vaccinations and tests conducted outside hospital systems may not be captured accurately in the EHR.

Lastly, cardiovascular diagnoses were based on ICD or SNONED codes without chart review confirmation, which can lead to misclassification. Also, the absence of family history data in our EHR limits understanding of the predisposition of patients to cardiovascular conditions due to familial trends.

In summary, this study shows a heightened risk of cardiovascular disease in children following SARS-CoV-2 infection, with similar risks observed in those with and without pre-existing congenital heart disease. Awareness of cardiovascular complications in the post-acute phase will improve timely diagnosis and treatment of these conditions.

## Data Availability

The data can be shared with the request to the RECOVER initiative (https://recovercovid.org/)

## Acknowledgments

This study is part of the NIH Researching COVID to Enhance Recovery (RECOVER) Initiative, which seeks to understand, treat, and prevent the post-acute sequelae of SARS-CoV-2 infection (PASC). For more information on RECOVER, visit https://recovercovid.org/.

We would like to thank the National Community Engagement Group (NCEG), all patients, caregivers, and community Representatives, and all the participants enrolled in the RECOVER Initiative. We would like to thank the patient representatives Megan Carmilani, Nick Guthe, and Leah Baucom for their helpful suggestions and comments.

## Supplemental Material

Protocol and Statistical Analysis Plan

Supplemental Methods/Dataset/Tables S1-S3/Figures S1_S15

